# Sex-Specific Diagnostic Subtypes in Adolescents Hospitalized for Substance Use Disorders Revealed by Transformer-Based Clustering

**DOI:** 10.1101/2025.08.06.25333108

**Authors:** Manuel Corpas, Hilario Blasco, Lucía Gallego, José Manuel Ramos-Rincón, Víctor Moreno-Torres, Vicente Soriano

## Abstract

Substance use disorders (SUD) are a leading cause of psychiatric hospitalization among adolescents, yet the underlying diagnostic profiles and comorbidities remain poorly characterized. Here, we applied a transformer-based language model to 4,849 hospital discharge records from adolescents (aged 11–18) admitted with mental health and SUD in Spain between 2016 and 2020. We generated dense clinical embeddings and identified sex-specific diagnostic clusters using unsupervised learning. Male subgroups were primarily defined by externalizing disorders such as conduct disorder, ADHD, and psychosis, often associated with cannabis and alcohol use. Female subgroups exhibited higher rates of borderline personality traits, mood instability, suicidal ideation, and stress-related conditions. A unified embedding space revealed a clear sex-based separation in diagnostic patterns. These findings highlight the potential of transformer-based embeddings to uncover latent clinical phenotypes and inform precision approaches to adolescent mental health care. These sex-specific subtypes could enhance early intervention strategies and support the development of tailored treatment programmes for youths with SUD.

## Introduction

Adolescence is a vulnerable developmental period when the initiation of drug use can precipitate or exacerbate a range of mental health problems. During the past two decades, nearly 6% of all hospitalizations in Spanish 11–18-year-olds have involved a psychiatric disorder, and substance use disorders (SUD) are the single most common category, accounting for about 40% of adolescent psychiatric admissions [1]. Co-occurrence of multiple mental disorders in youth is highly prevalent: individuals diagnosed with one psychiatric disorder often suffer or develop others [2]. This overlap is associated with worse clinical outcomes, including elevated premature mortality and a greater burden of disease [3].

Identifying distinct clusters of comorbid conditions can yield insights into shared etiological pathways and help tailor more effective, integrated interventions [4]. For example, grouping patients by common comorbidity profiles may enable specialized youth care programs (such as targeted services for adolescents with concurrent mood disorders and SUD) and ultimately improve outcomes by addressing more specifically the needs of each subgroup [5].

Unsupervised machine learning techniques like cluster analysis offer a data-driven approach to uncovering these multidimensional patterns of illness. Prior studies on multimorbidity have found reproducible groupings of diseases [6]. For instance, patients with psychiatric disorders tend to form their own cluster apart from those with predominantly physical ailments [6]. However, most clustering approaches in health research have been applied to *diseases* (identifying groups of correlated diagnoses), and relatively few studies have clustered *individual patients* based on a holistic view of all their comorbidities [6]. Clustering patients by their full multi-diagnosis profiles may improve our understanding of shared causes and prognoses and inform health service design. In the context of adolescents with SUD, such an approach may reveal clinically meaningful subgroups. Instances could include identifying a cluster of youth characterized by primary substance misuse with co-occurring behavioral problems, versus another cluster defined by mood/anxiety disorders with incidental substance use. Identification of these subpopulations can be critical for developing targeted prevention and treatment strategies.

Despite its promise, clustering heterogeneous mental health data poses significant challenges. Psychiatric disorders span diverse symptom domains and diagnostic categories, making co-occurrence patterns high dimensional and complex. Patients often carry long lists of ICD codes ranging from mood and anxiety disorders to psychotic, neurodevelopmental, and SUD, a level of heterogeneity that is difficult to capture with traditional one-hot encoded feature vectors. Moreover, some diagnoses are quite common (e.g. major depression) while others are rare. Naïve clustering can be dominated by prevalent conditions and may overlook subtler but clinically meaningful groupings. There is also no obvious *a priori* way to choose the number of clusters, since mental health diagnoses do not neatly fall into known subclasses. Finally, interpretability is a concern: any clustering solution must be explainable in clinical terms to be useful, yet making sense of the relationships among dozens of co-occurring disorders is not straightforward.

Recent advances in natural language processing offer a potential remedy to several of these difficulties. Large language models (LLMs) are neural networks pre-trained on vast text corpora. They have been adapted to biomedical applications including representation learning for electronic health records. By encoding the *semantic* meaning of clinical codes and descriptions, LLMs can transform a list of a patient’s diagnoses into a dense vector embedding that reflects underlying clinical similarity. For example, transformer-based models such as BEHRT [7] and Med-BERT [8] have been used to learn patient representations from longitudinal EHR sequences, effectively placing semantically related diagnoses closer together in the model’s latent space. An LLM can thus position, say, *generalized anxiety* nearer to *major depression* than to *autism* or *schizophrenia* in the vector space. Clustering in this embedding space is expected to yield more coherent and clinically sensible groups of patients compared to using raw code-based distances. Importantly, coupling LLM-derived features with rigorous clustering techniques promises not only to improve the detection of true subgroups, but also to preserve interpretability by allowing us to trace each cluster back to its defining clinical features. To date, however, such transformer-based representation learning approaches have not been widely applied to cluster psychiatric comorbidity patterns, especially in adolescent populations.

Our study focuses on Spanish adolescents hospitalized with drug-related mental health disorders. By applying an LLM-based clustering approach we aim at identifying distinct phenotypic subgroups. We utilize a large nationwide public hospital discharge database [9] to capture real-world co-occurrence of mental and behavioral disorders in youths with substance use. We then employ a transformer model [10] to embed each patient’s diagnostic profile into a vector representation, and perform robust unsupervised clustering on these embeddings to uncover natural groupings of patients. This approach marries state-of-the-art representation learning with data-driven clustering to identify clinically meaningful clusters of individuals with multiple mental health conditions.

## Methods

### Spanish National Registry of Hospital Discharges (RAE-CMBD)

The data source for this study was the Spanish National Registry of Hospital Discharges (RAE-CMBD), an administrative database that compiles information on virtually all hospital discharges in Spain [11,12]. Established via the Minimum Basic Data Set (MBDS) in the late 1980s, the RAE-CMBD requires mandatory reporting from all public hospitals (and since 2005, private hospitals as well), resulting in a nationwide registry that captures over 90% of acute-care hospitalizations [12]. Given Spain’s universal health coverage, which insures approximately 99.5% of the population [13], the RAE-CMBD provides a comprehensive representation of hospital admissions across the entire nation. For each recorded discharge, the registry includes detailed demographic and clinical information: patient characteristics (e.g., age and sex), the primary diagnosis (and any secondary diagnoses) at hospital discharge, procedures performed, and health outcomes such as length of stay and in-hospital mortality [12].

The RAE-CMBD has been extensively utilized for healthcare planning, resource allocation, and epidemiological research [11]. Its rich dataset has informed budget distribution and the organization of health services, for example, guiding hospital bed availability and specialized staff requirements [11]. Moreover, RAE-CMBD has supported numerous population-wide studies across diverse disease areas from infectious diseases (e.g. HIV, viral hepatitis, COVID-19) to chronic illnesses like cardiovascular and respiratory conditions [14–16]. This underscores its value in public health surveillance and policy-making.

During a full 22-year observation window (2000–2021), our RAE-CMBD data dump captured 2,015,589 hospitalizations of adolescents with mental disorders in Spain [11]. Within this cohort, 118,609 (5.9%) involved at least one diagnosed psychiatric condition [11]. Notably, while the total volume of adolescent hospitalizations steadily decreased over time, the proportion of those linked to mental disorders rose from 3.9% in 2000 to 9.5% in 202, illustrating an increasing burden of psychiatric conditions among Spanish youth [11].

For this analysis, we selected all adolescent hospital discharge records from the RAE- CMBD involving SUD, spanning January 1, 2016, through 15^th^ of March, 2020. This narrower window offers a coherent frame of reference for detecting changes in adolescent mental health admissions leading up to the COVID-19 pandemic, without the confounding influence of pandemic-related shifts that became pronounced after March 15^th^ 2020. All diagnoses and procedures in the database were coded using the International Classification of Diseases (ICD) system; specifically, the Tenth Revision (ICD-10-CM) was employed for discharges throughout this period [17].

Within the collected records, we identified those involving mental health conditions, focusing on 19 distinct mental disorders documented in the discharge diagnoses (**Supplementary Table S1**). These encompassed a broad range of psychiatric conditions in adolescents (for example, mood, anxiety, eating, and developmental disorders, among others), ensuring that our analysis captured the spectrum of major mental health-related hospitalizations in this age group. For each hospitalization meeting these criteria, we examined the associated clinical data and outcome measures. In particular, we analyzed the hospital outcomes of interest (including the length of hospital stay and in-hospital mortality) for adolescents with mental disorder diagnoses.

### Data Source and Pre-Processing

We focused on adolescent patients (ages 11-18) admitted between 1^st^ of January, 2016 and 15^th^ of March, 2020 with SUD diagnoses, identified using ICD-10-CM codes F11-F16 and F18-F19. Their main features are summarized in **Supplementary Table 2**. Cases with isolated alcohol or tobacco use disorders (with no mental disease cooccurences) were excluded. We also excluded patients whose primary diagnosis was not within the F code subset of ICD-10. In order to assess and report health data we followed RECORD guidelines [18].

Each hospitalization record contained up to 20 ICD-10-CM diagnoses, including a primary diagnosis and additional comorbid conditions. To construct a comprehensive patient profile, we aggregated all admissions associated with each anonymized patient ID, capturing the full spectrum of diagnoses assigned during adolescence.

We then split the dataset by sex, creating two subsets: one for male patients (sex = 1) containing 2,467 admission records and one for female patients (sex = 2) containing 2,382. For each subset, we prepared the diagnostic text for language-model analysis by mapping each ICD-10 code to a standardized descriptor (for example, F11.2 → “opioid dependence”), or “NOCODE” if no descriptor was available. These descriptors were concatenated into a single diagnostic sentence per patient. In all analyses, we stratified by the number of diagnoses and retained 90% of each sex-specific subset for training and 10% for validation, ensuring even representation of patients with multiple diagnoses.

### Language Model Training and Embedding Generation

To transform diagnostic text into numerical representations suitable for clustering, we employed DeBERTa-v3 [6], a transformer-based language model. We fine-tuned the model separately on the male and female subsets. This sex-stratified fine-tuning was necessary because male and female adolescents exhibit distinct patterns of psychiatric comorbidity [19–21]. Training separate models ensured that these differences were preserved in the embedding space, improving the ability of the clustering algorithm to detect meaningful, sex-specific diagnostic subtypes.

#### 1. Masked Language Modeling (MLM) Pretraining

For each sex-specific subset, we adapted DeBERTa-v3 to the domain of adolescent mental health by training it with masked language modeling (MLM). In this step, 15% of tokens in each diagnostic sentence were randomly masked, and the model was trained to predict the missing tokens based on the surrounding context. This procedure improved the model’s capacity to capture psychiatric terminology and patterns of co-occurring diagnoses. Each MLM phase was run for 5 epochs, using a batch size of 16 and an initial learning rate of 5×10⁻⁵.

#### 2. Contrastive Fine-Tuning for Embedding Stability

Within each sex-specific dataset, we next applied a contrastive learning objective to stabilize the learned representations. Each patient’s diagnostic sentence was augmented into two perturbed versions (by shuffling or omitting a small number of diagnoses), treated as “positive” pairs, while texts from different patients were considered “negative” pairs. A contrastive loss encouraged embeddings of positive pairs to remain close, while pushing apart embeddings derived from dissimilar profiles.

This second phase used 5 epochs, a batch size of 16, and an initial learning rate of 1×10⁻⁵.

Following contrastive fine-tuning, each male or female patient was assigned a 768- dimensional embedding by passing the diagnostic text through the corresponding sex-specific model and applying mean pooling over the final hidden-layer tokens. We thus obtained one set of embeddings for males and a separate set for females.

### Clustering Analysis

We performed unsupervised clustering separately on the male and female embeddings, aiming to discover distinct diagnostic subgroups within each sex:

#### 1. Optimal Cluster Determination via Bootstrapping

To determine the optimal number of clusters (K), we performed a bootstrap-based silhouette analysis for both the male and female embedding sets. In each iteration, a random subset of patient embeddings was clustered, and silhouette scores were computed to assess cluster coherence. Candidate values of K from 2 to 10 were tested, and the K with the highest average silhouette score across bootstraps was selected to balance granularity and stability.

#### 2. K-Means Clustering

Having identified the optimal K, we performed k-means clustering on the respective full embedding set (males or females). This generated two final sets of clusters: one exclusively for male patients and one for female patients.

#### 3. Cluster Characterization via c-DF–IPF

To reveal the most characteristic diagnoses in each male-specific or female-specific cluster, we computed cluster-based disease frequency-inverse patient frequency (c- DF-IPF) scores. Inspired by TF-IDF, c-DF-IPF measures how frequently a diagnosis appears in a particular cluster relative to its overall prevalence in that subset. Specifically:

1. **Disease Frequency (DF):** The proportion of patients in a cluster assigned a given ICD-10 code.
2. **Inverse Patient Frequency (IPF):** The logarithm of the total patient count (in that subset) divided by the number of patients diagnosed with the code.
3. **c-DF-IPF Score:** The product of DF and IPF, emphasizing diagnoses that are both common within a cluster yet relatively rare across the rest of the same sex.

The top 5 highest-scoring ICD-10 codes in each cluster were used to interpret and label the resulting male and female clusters. Example clusters included “Mood/Anxiety with Substance Use” and “Psychosis with Dual Diagnosis”, reflecting patterns in adolescent mental health disorders.

### Visualization and Interpretation of Clusters

After establishing male- and female-specific clusters via the silhouette-based K selection, k-means clustering, and c-DF-IPF scoring (as described above), we conducted a post-hoc visualization step to more intuitively compare and interpret the resulting subgroups. Specifically, we sought to (a) map each existing cluster onto a concise 2D projection for visual inspection, and (b) derive succinct “dominant semantic terms” that characterize each cluster’s core diagnostic themes.

#### Cluster-Level Embeddings and 2D Projection

Although clustering had already been performed separately for males and females, we combined all patient records (male and female) into a unified embedding space to facilitate comparisons across the sexes. For each patient record, we converted the set of ICD-10 diagnoses into textual descriptors removing dots, mapping codes to short descriptions where possible, and replacing inter-word spaces with underscores (e.g., Cannabis abuse, uncomplicated -> cannabis_abuse,_uncomplicated).

We then applied a standard TF-IDF vectorization to all textual documents (one per patient), producing a high-dimensional representation in which each token reflects a particular ICD descriptor.

Because each record was already assigned to one of the male or female clusters, we grouped the TF-IDF vectors by cluster and averaged the embeddings within each cluster. This yielded a single cluster-level embedding per subgroup, one point representing all patients in that cluster.

To visualize these cluster-level embeddings, we performed two 2D projections (using PCA and UMAP). In the resulting scatter plots, each point corresponds to a previously identified cluster. This approach highlights how each cluster, generated by the earlier k-means procedure, lies relative to other subgroups in the semantic space of diagnoses. Clusters lying near each other in 2D typically share similar high-level patterns of diagnoses.

#### Dominant Semantic Terms

To provide interpretable summaries of each cluster’s diagnostic content beyond the c- DF-IPF codes, we further examined the TF-IDF features. For each cluster, we collected all TF-IDF vectors from the individual patients assigned to that subgroup. We then computed the mean TF-IDF values across all patients in the cluster, essentially the same step used to generate the cluster-level embedding above. Finally, we identified the highest-valued tokens (e.g., *“cannabis_abuse,_uncomplicated”* or *“brief_psychotic_disorder”*). Because these tokens reflect descriptors that are both frequent and distinctive, they form a concise “dominant semantic description” of the subgroup. We extracted the top five tokens to characterize each cluster.

These additional TF-IDF-based terms serve as a complementary interpretation to the c-DF-IPF codes. While c-DF-IPF highlights the most characteristic ICD-10 diagnoses in terms of cluster frequency vs. broader prevalence, the TF-IDF tokens reveal the most discriminative descriptors from a text-based perspective. These visual 2D projections and the cluster-level dominant terms may help clarify how previously discovered male and female clusters differ in their diagnostic profiles, providing an accessible snapshot of key conditions that define each subgroup.

### Ethics

The Spanish National Registry of Hospital Discharges (RAE-CMBD) database used for this analysis has been provided by the Spanish Ministry of Health and is available upon request. All potential patient identifiers have been removed from the analysis. According to Spanish law, patient informed consent is not needed for this analysis. The study was approved by the Clinical Research Ethics Committee of the Universidad Internacional de La Rioja (UNIR) (ref. PI-022/2023; 4/27/2023). All research has been carried out following ethical standards as defined by the revised Declaration of Helsinki, 2013.

## Results

### Overall Cohort Distribution

Of the 4,849 hospital discharge records analysed, 2,467 were associated with male and 2,382 with female adolescents. Both sexes showed a different year-by-year pattern, peaking in 2019 and decreasing thereafter (**Figure 1**). This difference in distribution was statistically significant (χ² (4) = 13.589, *p* = 0.00873). Year 2020 ended on March 15^th^, which means that only two and a half months were taken into consideration.

**Figure 1.**
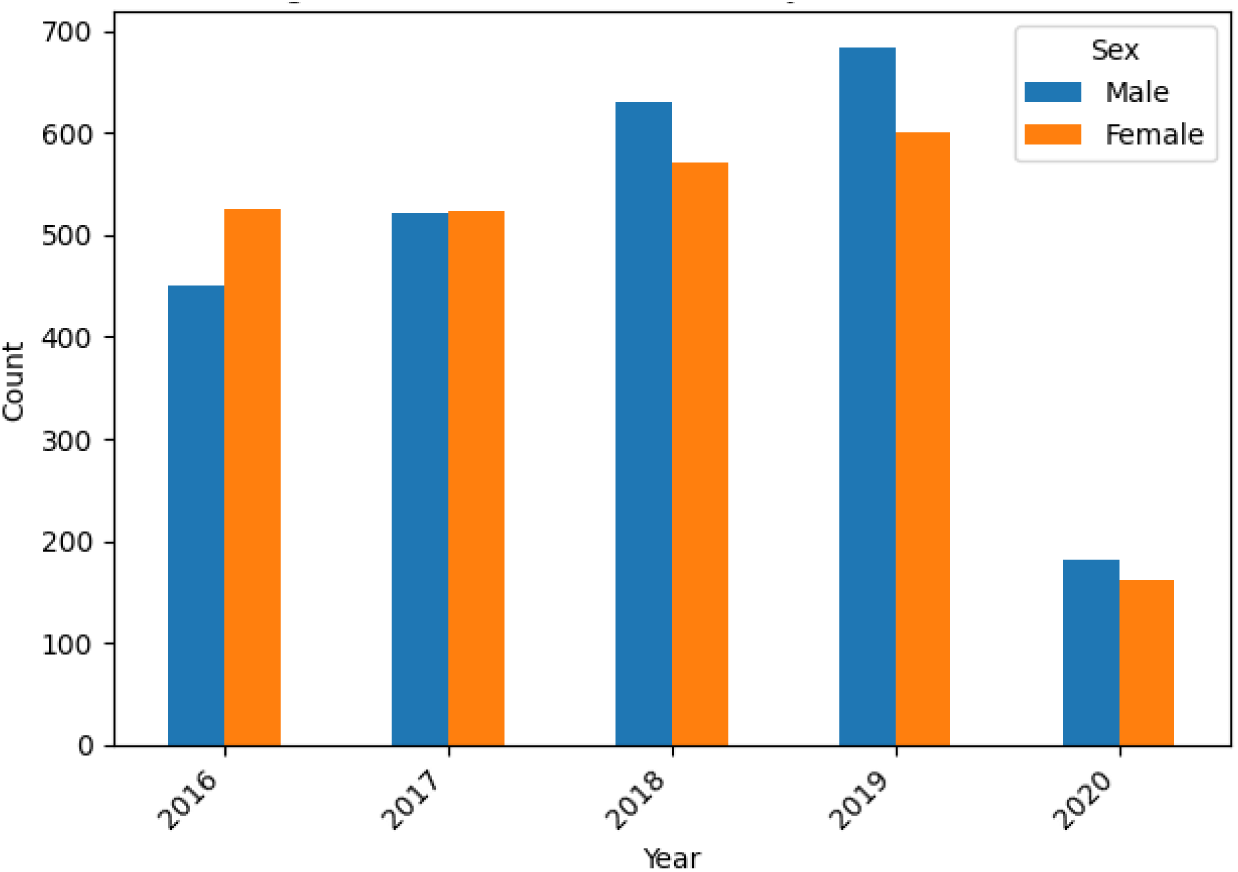
Year-by-year distribution of adolescent hospital admissions with SUD and mental health comorbidities (2016-2020) among **male** and **female** patients. Each bar represents the total number of discharges recorded for that year in the respective sex group (patient data collection stops on March 15^th^ 2020, hence why the steep decline for that year). Both distributions peak in 2019 and decline thereafter, a chi-square test revealed statistically significant difference in overall yearly patterns between males and females (χ² (4) = 13.589, *p* = 0.00873).

### Patient Diagnostic Profiles

A striking contrast emerged when comparing main diagnosis between male and female groups (**Figure 2**). Males frequently presented with F29 (unspecified psychosis), F23 (acute and transient psychotic disorders), and F91.9 (unspecified conduct disorder). This pattern underscores a notable presence of psychotic and disruptive/conduct- related diagnoses.

**Figure 2.**
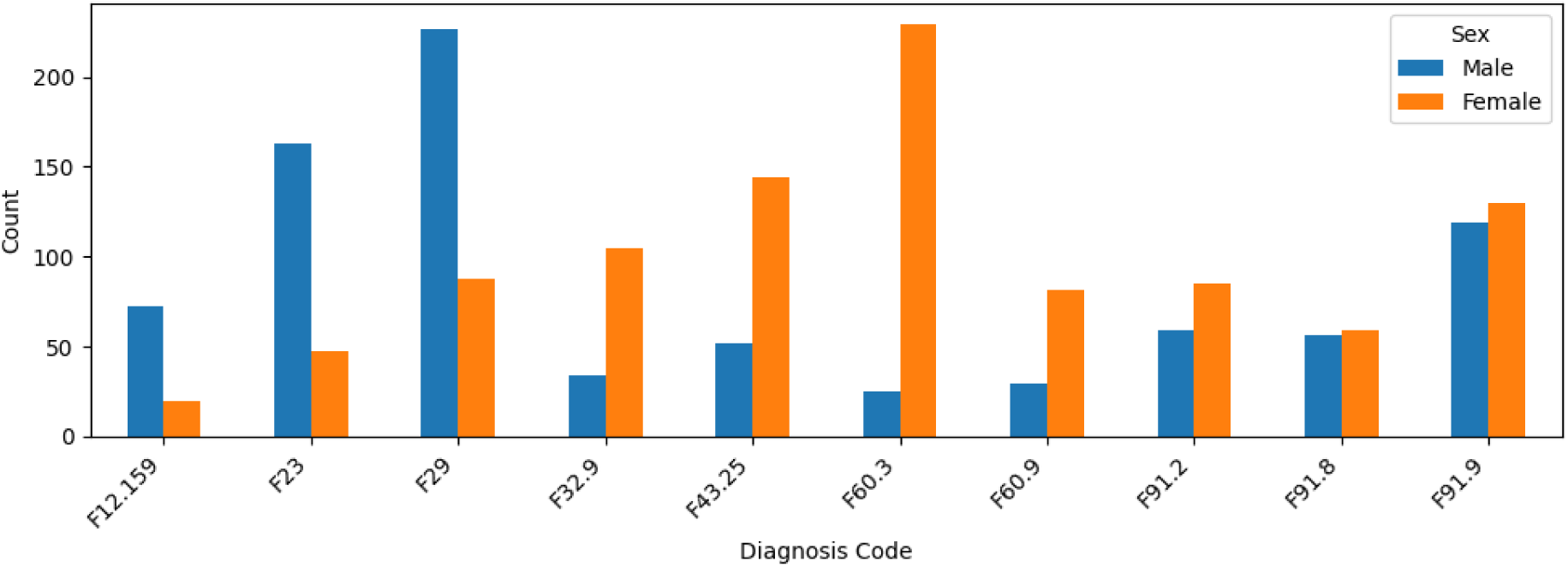
Top 10 principal diagnoses observed in **male** adolescents (top) versus **female** adolescents (bottom). Each bar indicates the total number of patients assigned the corresponding ICD-10 code. Compared with females, males presented more frequently with psychotic and disruptive/conduct-related conditions (e.g., F29, F23, and F91.9). In contrast, females showed higher rates of borderline personality disorder (F60.3) and other stress-related diagnoses (F43.25).

In contrast, females showed higher proportions of F60.3 (borderline personality disorder), F43.25 (adjustment disorder with mixed anxiety and depressed mood) and F91.9 (unspecified conduct disorder). Notably, the female subset included more personality disorders and stress-related diagnoses.

When the diagnostic analysis was narrowed to the top 20 diagnoses (plus an “OTHER” category), a chi-square test revealed a marked and highly significant difference between sexes (χ² (20) = 3.731, *p* = 1.76e-117). This finding indicates that the overall diagnostic landscape for adolescent inpatients with SUD differs substantially between males and females, with males more likely to be admitted for severe psychotic or conduct-related presentations, and females more frequently hospitalized for personality-related disorders, or mood/anxiety disorders, roughly corresponding to classical externalizing and internalizing mental illnesses, respectively.

### Length of Stay and Severity

Although males demonstrated a longer hospital stay on average (median = 10 days, IQR = 5-17) relative to females (median = 9 days, IQR = 5-16) (**Figure 3**), this one-day gap in median stay was not statistically significant (U = 2983361.5, p-value=0.354).

**Figure 3.**
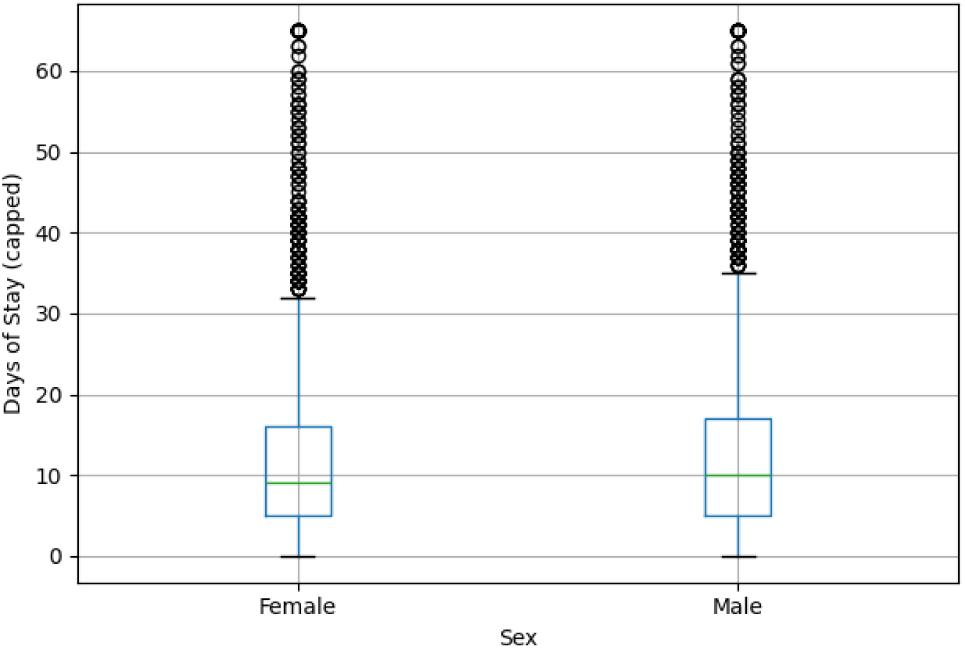
Box plots comparing the length of hospital stay (Days of Stay) in **females** (left) and **males** (right). Data are capped at the 99th percentile to limit extreme outliers. The horizontal line within each box denotes the median, with the box boundaries representing the interquartile range (IQR), and individual circles indicating data points beyond the upper whisker. Males exhibit a higher median stay whereas females show slightly shorter admissions. Difference in stays between males and females is not statistically significant (p-value=0.354).

In line with this, the severity level (“APR Severity Level”) distribution did not vary significantly between sexes (χ^2^(3) = 3.731, p-value=0.292) (**Figure 4**).

**Figure 4.**
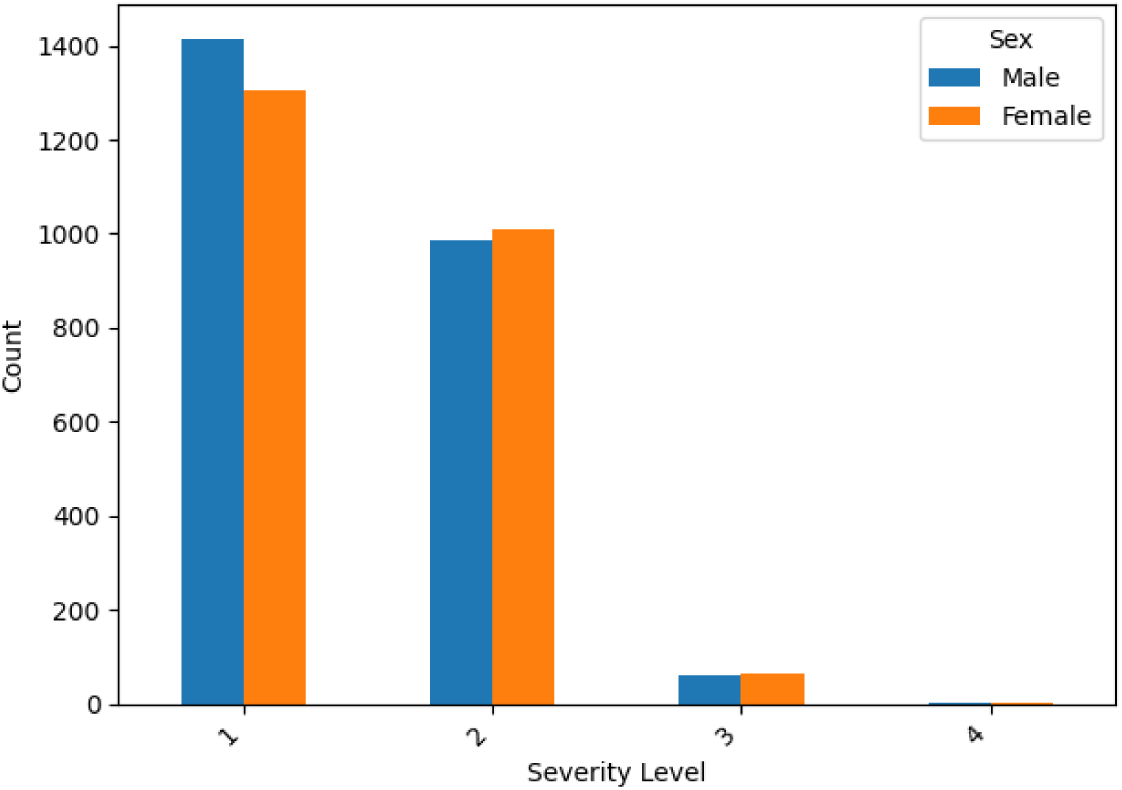
Distribution of APR severity levels (0-4) for males versus females. Each bar indicates the number of patients classified at a particular severity category. The overall disparity in severity distribution between the sexes was not significant (χ^2^(3) = 3.731, p-value=0.292).

### Resource Utilization

The hospital cost analysis underscores the distinction between the sexes. Males incurred a higher median cost (4,466.63 euros vs. 4,317.17 for females), and this disparity was highly significant (U = 3,226,352.0, p-value=3.35e-09). The findings imply that although male adolescents do not only stay longer in the hospital, they require more resources and potentially more complex interventions, in line with their elevated prevalence of psychotic and disruptive disorders.

### Distribution and Structure of Male and Female Diagnostic Clusters

Using the unified TF-IDF embedding space, we analyzed the relative positions of male and female diagnostic clusters derived from transformer-based embeddings and k-means clustering. **Figure 5A** displays the PCA projection of the cluster centroids, while **Figure 5B** provides the corresponding UMAP projection, highlighting spatial patterns that reflect diagnostic similarity and sex-specific separation.

**Figure 5.**
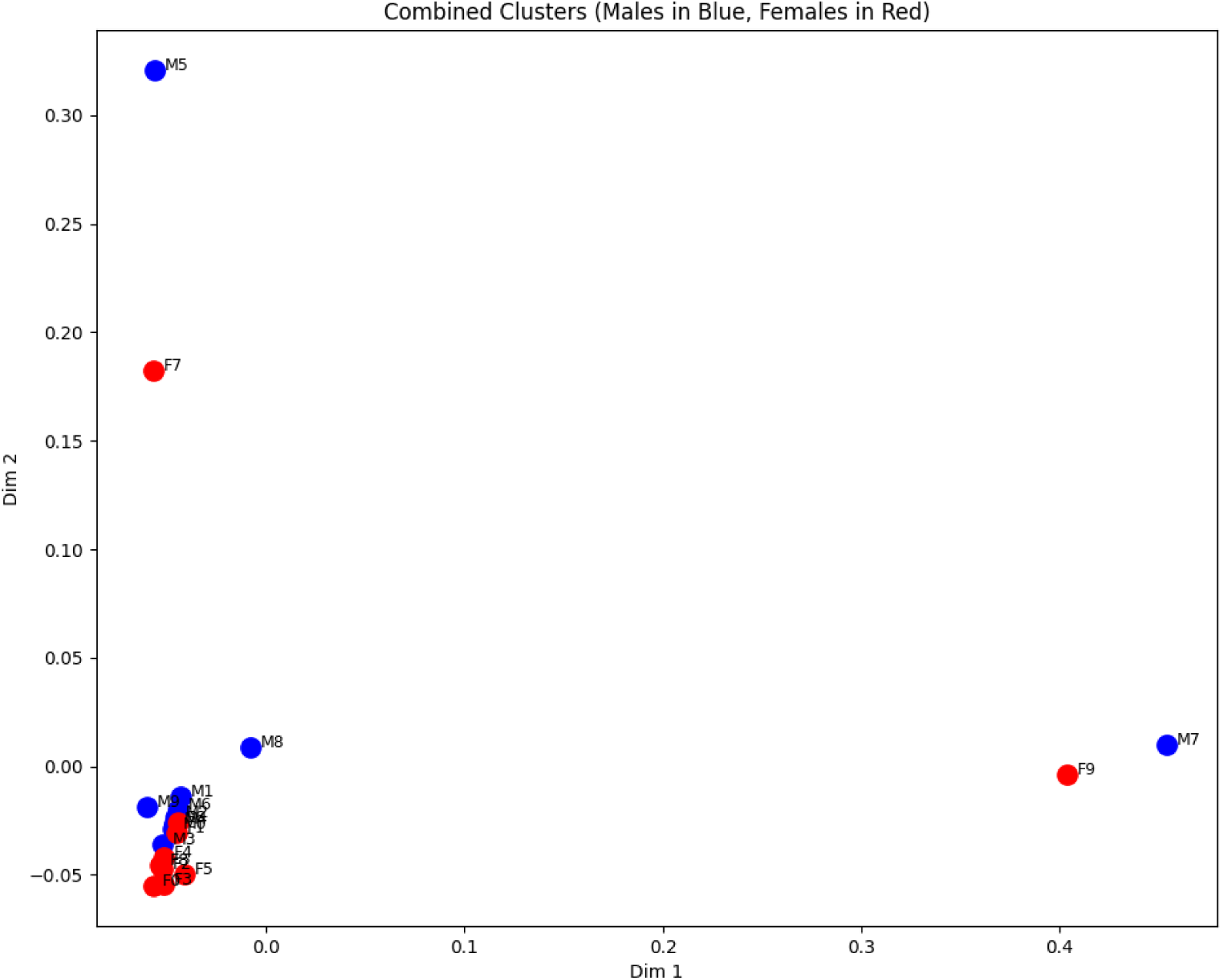
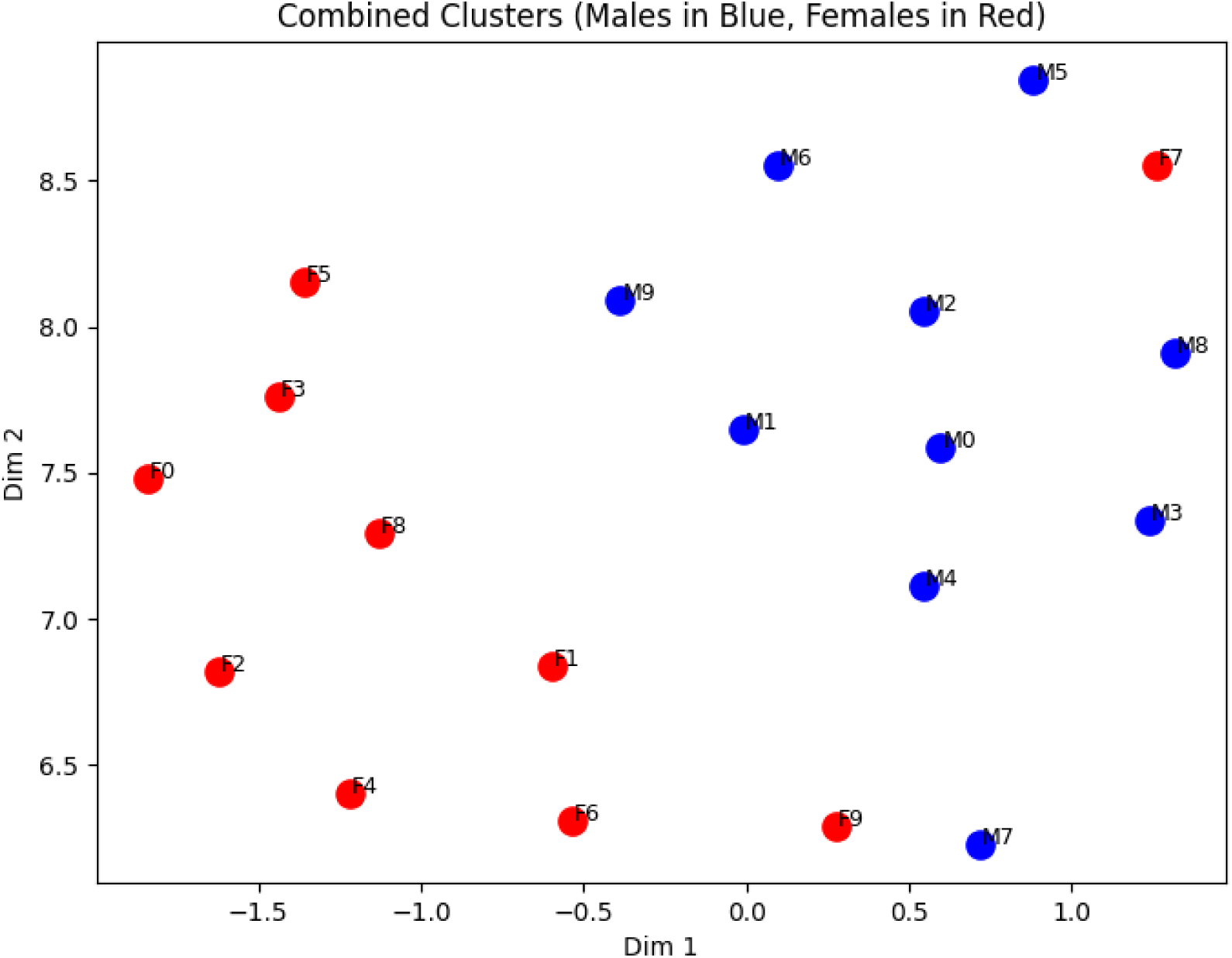
Combined PCA Projection of Male and Female Clusters. This figure illustrates both shared and sex-specific structure in the distribution of adolescent psychiatric diagnoses, supporting the presence of distinct phenotypic subgroups among hospitalised youth. (**A**) Two-dimensional PCA projection of diagnostic cluster centroids derived from adolescent hospital discharge records (n = 4,849). Each point represents the average TF-IDF embedding of a k-means cluster. Blue dots indicate male clusters (M0-M9), and red dots indicate female clusters (F0-F9). Axes correspond to the first two principal components. Clusters closer together share more similar diagnostic profiles. Outliers such as M5 (psychosis with intellectual disability) and F7 (perinatal complications and borderline traits) occupy the upper periphery, while M7 and F9 separate along the horizontal axis due to distinctive substance- psychosis combinations. (**B**) UMAP projection of the same cluster centroids. This non-linear embedding reveals a clear sex-based separation along the horizontal axis: male clusters align on the right, and female clusters on the left, with minimal overlap. Vertical positioning reflects intra-sex heterogeneity, distinguishing externalising clusters from those defined by psychosis, borderline traits, or developmental delay.

In the PCA space (**Figure 5A**), most male (blue) and female (red) clusters aggregate tightly near the origin, indicating a central hub of shared diagnostic features, particularly for those clusters dominated by substance use, conduct disorder, and personality traits. However, four clusters exhibit clear separation along the principal components. On the positive side of PC1, M7 (cannabis use and ADHD) and F9 (female psychosis with substance abuse) form a mirrored outlier pair, while M5 (psychosis plus intellectual disability) and F7 (female perinatal cases with borderline personality disorder) diverge along PC2, occupying the upper extremes. These positions suggest the existence of rarer phenotypes with distinctive comorbidity patterns, less commonly shared across sexes.

In contrast, the UMAP projection (**Figure 5B**) reveals a clear horizontal dichotomy between male and female clusters. All male clusters lie to the right (positive *Dim 1*), and all female clusters to the left, with only two exceptions near the midline: M9, a male group with cannabis-associated psychosis, and F1, a female cluster featuring surgical and ENT codes alongside substance use. This suggests that while PCA captures broad variance, UMAP uncovers non-linear separability that more accurately reflects underlying sex-specific diagnostic structure.

Within each sex-defined region, UMAP also preserved meaningful vertical differentiation. Male clusters ascended from externalising profiles (e.g., M0–M4) toward more psychosis- or intellectual disability-oriented presentations (e.g., M5–M7), while female clusters ranged from borderline and mood disorders (e.g., F0–F4) to more complex and rare profiles involving psychosis or developmental delays (e.g., F7, F9).

This topography was consistent with cluster content derived from c-DF–IPF and semantic analysis. For instance, M7, located at the far right of both projections, was defined by high rates of unspecified psychosis (F29), cannabis dependence, and nicotine use. Its closest female counterpart in terms of psychotic orientation, F9, also featured heavy psychosis coding but was slightly more centrally located, likely due to its additional complexity (e.g., comorbid infections, substance use). At the opposite end, F0, F1, and F4 formed a compact group of cannabis-abusing females with borderline traits, closely aligned with M0 and M4, two male clusters dominated by cannabis, conduct, and alcohol use disorders.

Whereas PCA reveals a partially overlapping core structure dominated by SUD, UMAP exposes a fundamental bifurcation along sex lines, with diagnostic diversity preserved within each sex. These findings suggest that while adolescents of both sexes hospitalized with SUD present broadly similar conditions, their patterns of comorbidity form distinct geometries in embedding space. These geometries may require tailored approaches in clinical care and resource allocation. **Suplemmentary Table S3** summarizes the characteristics of all 20 diagnostic clusters (F0-F9 for females; M0-M9 for males).

## Discussion

This study applied a transformer-based representation learning approach to characterize sex-specific diagnostic subgroups among adolescents hospitalized for substance use disorders (SUD). By embedding diagnostic code profiles into a dense semantic space and applying unsupervised clustering, we identified distinct patterns of psychiatric comorbidity that differed substantially between male and female youths.

Our results show that male adolescents were predominantly grouped into clusters characterised by externalizing conditions, including conduct disorder, attention-deficit/hyperactivity disorder (ADHD), and psychotic disorders. These clusters often relate to cannabis and alcohol misuse. By contrast, female adolescents exhibited a diagnostic landscape dominated by internalizing symptoms, notably borderline personality disorder traits, adjustment disorders, mood instability, and suicidality.

These findings align with broader epidemiological data suggesting divergent developmental pathways between male and female adolescents in the emergence of psychiatric disorders [19,22,23]. Externalizing disorders typically onset earlier in males and are associated with impulsivity, sensation-seeking, and behavioral dysregulation [24,25], whereas internalizing symptoms such as mood and anxiety disorders become more prevalent in females during adolescence, potentially influenced by hormonal, neurodevelopmental, and psychosocial changes [26,27]. Our study expands upon these observations by applying a high-dimensional clustering framework that makes these sex-specific distinctions at the level of hospitalized patients with complex dual diagnoses.

Importantly, the diagnostic embeddings captured subtle co-occurrence patterns that may reflect shared vulnerability factors or clinical trajectories. For instance, the association between cannabis use and psychosis was prominent among male adolescents, consistent with evidence suggesting that early cannabis exposure, particularly potent strains, can precipitate psychotic symptoms in genetically or developmentally susceptible individuals [28]. This underscores the need for early substance use screening and psychosis risk assessment among male youths presenting with disruptive behaviour or SUD.

In females, the frequent co-occurrence of substance use with borderline personality traits and mood disorders suggests a different clinical profile where substance use may serve as a maladaptive coping mechanism for underlying affective dysregulation or trauma. The observed clustering of perinatal complications and psychiatric symptoms in some female subgroups also highlights the potential intersection between reproductive health and mental health care during adolescence, a critical but often overlooked aspect of youth psychiatric services [29].

Beyond confirming known sex differences, our approach revealed novel diagnostic constellations that may guide more targeted interventions. For example, clusters combining intellectual disability with psychosis or substance use suggest a high-need subgroup requiring integrated developmental and psychiatric care. Similarly, clusters linking substance misuse with acute somatic conditions (e.g., respiratory or ENT issues) point to the importance of liaison services bridging emergency medicine and psychiatry for adolescents with complex presentations.

From a methodological perspective, the use of transformer-derived embeddings offers several advantages over traditional clustering based on raw diagnostic codes. First, it allows a more nuanced representation of clinical similarity, where semantically related conditions are placed closer together in latent space, enhancing cluster coherence. Second, it mitigates issues related to data sparsity and rare diagnoses, improving the detection of smaller but clinically meaningful subgroups. Finally, by fine-tuning the language model on sex-stratified data, we were able to amplify diagnostic patterns specific to each sex, although this approach also introduces some limitations regarding direct cross-sex comparison.

### Clinical Implications

Our findings have tangible clinical relevance. Adolescent mental health services should consider sex-specific diagnostic architectures when designing prevention and treatment programmes. For male adolescents with SUD, interventions might benefit from integrating conduct disorder management, psychosis early detection, and substance misuse treatment within a unified care pathway. For female adolescents, trauma-informed care models that address emotion regulation, interpersonal difficulties, and comorbid mood disorders may be more appropriate.

Moreover, the identification of latent diagnostic subgroups supports the move toward precision psychiatry in adolescent populations. Embedding-based phenotyping could complement clinical assessment by offering data-driven insights into patients’ risk profiles, comorbidity patterns, and potential care trajectories. In practical terms, embedding-based clustering could support triage systems by flagging high-risk profiles (e.g., psychosis with intellectual disability) for more intensive case management, or guide referral decisions by matching patients to tailored care pathways. These phenotypic embeddings may also serve as inputs to predictive models for relapse, treatment response, or readmission, offering a complementary layer to existing diagnostic assessment tools.

### Biological and Psychosocial Mechanisms

The sex differences observed likely arise from an interplay of biological and psychosocial factors. Neurodevelopmental studies have shown sex-specific trajectories in brain maturation, particularly in regions implicated in impulse control, emotional regulation, and reward processing [30–32]. Hormonal changes during puberty, including shifts in oestrogen and testosterone levels, may further modulate vulnerability to psychiatric disorders and substance use [33,34]. Additionally, gender-related social norms and expectations may influence help-seeking behaviors, diagnosis assignment, and hospitalization thresholds, potentially shaping the patterns observed in administrative data. Future research should aim to disentangle these biological and psychosocial contributions to better tailor interventions.

Although we stratified the analysis by binary sex, we acknowledge that diagnostic coding systems and hospital records often conflate sex and gender, potentially masking experiences of gender-diverse adolescents. The absence of data on gender identity, ethnicity, or socioeconomic status limits our ability to examine intersecting dimensions of mental health inequity. Moreover, differential diagnosis thresholds, clinician bias, and healthcare access disparities may shape the sex-specific patterns observed. Future work should incorporate gender diversity and intersectional factors to more comprehensively address mental health equity in youth populations.

## Limitations and Future Directions

Several limitations warrant consideration. Our analysis relied on hospital discharge data, which may underrepresent adolescents with less severe or subclinical presentations managed in outpatient settings. The use of ICD-10 coding, while standardized, may not fully capture the complexity of psychiatric comorbidity, and diagnostic practices may vary across clinicians and institutions. Although our sample was nationally representative of Spain, cultural factors may limit generalizability to other populations. Additionally, by stratifying embeddings by sex prior to clustering, we enhanced sensitivity to sex differences but limited direct cross-sex comparability at the embedding level.

Future studies should validate these findings in independent cohorts, including outpatient populations, and explore whether embedding-derived phenotypes predict clinical outcomes such as relapse, hospital readmission, or treatment response. Longitudinal applications of transformer models could also examine how diagnostic profiles evolve over time in adolescents with dual diagnoses. Finally, expanding analyses to include gender identity and socio-environmental variables would provide a more comprehensive understanding of diagnostic subtypes beyond binary sex categories.

## Conclusions

This study shows that transformer-based clinical embeddings can uncover clinically meaningful, sex-specific diagnostic subgroups among adolescents with mental health conditions hospitalized for substance use disorders. Male adolescents predominantly exhibited externalizing profiles, with high rates of conduct disorder, ADHD, and psychosis, often in the context of cannabis and alcohol misuse. Female adolescents, by contrast, were more frequently characterized by internalizing symptoms, including borderline personality traits, mood instability, stress-related disorders, and suicidal ideation.

By embedding complex diagnostic profiles into a dense, semantically informed space, we were able to identify latent phenotypic structures that traditional clustering methods may fail to detect. These findings underscore the value of representation learning approaches in adolescent mental health research, offering new tools to refine diagnostic classification and uncover hidden patterns of comorbidity.

Clinically, the identification of distinct, sex-specific subgroups suggests that interventions for adolescents with SUD should be tailored to their broader psychiatric context. For male adolescents, early screening for psychosis and disruptive behavior disorders should be prioritized, whereas for females, trauma-informed, mood-stabilization, and emotion-regulation approaches may yield better outcomes. Resource allocation, care planning, and clinical guidelines in youth mental health services could benefit from incorporating these data-driven phenotypic insights.

Beyond immediate clinical applications, this work highlights the broader potential of embedding-based phenotyping to advance precision psychiatry in youth populations. Future studies should validate these patterns in diverse settings and investigate their predictive value for treatment outcomes and long-term trajectories. Expanding analyses to include gender diversity, socioeconomic context, and longitudinal follow-up will be critical to further tailoring mental health services to the nuanced needs of adolescents with complex dual diagnoses.

Given the use of nationwide public hospital data, these findings may also inform Spanish adolescent mental health policy and resource planning. Stratifying care models by sex-specific comorbidity patterns could thus improve early detection strategies and optimize the allocation of specialised services, particularly in psychiatric emergency and dual diagnosis care settings.

## Code Availability

All the data and code developed for this research are publicly available in the GitHub repository https://github.com/manuelcorpas/LLM-Mental-Health-Clustering. The repository provides Python scripts for data preprocessing, fine-tuning DeBERTa-v3, contrastive embedding, unsupervised clustering, and PCA/UMAP visualization, along with instructions for replicating each step. The data for males and females are available as CSV files with the corresponding predicted cluster pipeline result. Licensed under the MIT License, the code may be freely used and adapted for academic or other purposes, thereby promoting reproducible research and collaborative improvement of the pipeline.

## Supplementary Materials

Within the collected records from the Spanish National Registry of Hospital Discharges (RAE-CMBD), we identified hospital admissions that included a mental health condition, focusing on 19 distinct mental disorders (**Supplementary Table S1**). These diagnoses spanned a wide spectrum of psychiatric conditions in adolescents (including mood, anxiety, eating, and developmental disorders) thereby ensuring a comprehensive representation of mental health–related hospitalizations in this age group. These data have been previously reported in [11].

**Supplementary Table S1.** Stratification of 118,609 adolescent (ages 11–18) hospitalizations with mental disorders recorded in Spain from 2000 to 2021, by sex (male = 44.9%, female = 55.1%). “Total Hospitalizations” indicates the absolute count of admissions associated with each mental disorder; “Rate (%)” is the proportion out of all mental disorder admissions. “Male (%)” and “Female (%)” columns represent the sex distribution within each diagnostic category. *p*-values refer to comparisons of male vs. female proportions for each specific diagnosis (chi-square test). Note that “Substance use disorders (others than alcohol)” excludes primary alcohol use disorders, which appear as a separate category.

| Diagnosis | Total Hospitalizations | Rate (%) | Male (%) | Female (%) | p-value |
| --- | --- | --- | --- | --- | --- |
| Substance use disorders (others than alcohol) | 44,933 | 37.9 | 53.7 | 46.3 | < 0.0001 |
| Anorexia nervosa | 15,338 | 12.9 | 10.0 | 90.0 | < 0.0001 |
| Intellectual disability | 11,640 | 9.8 | 58.9 | 41.1 | < 0.0001 |
| Attention deficit hyperactivity disorder (ADHD) | 10,292 | 8.7 | 72.6 | 27.4 | < 0.0001 |
| Major depressive disorder | 9,881 | 8.3 | 25.7 | 74.3 | < 0.0001 |
| Other mood disorders | 9,822 | 8.3 | 38.5 | 61.5 | < 0.0001 |
| Posttraumatic stress disorder (PTSD) | 8,562 | 7.2 | 33.7 | 66.3 | < 0.0001 |
| Anxiety | 7,503 | 6.3 | 27.2 | 72.8 | < 0.0001 |
| Autism spectrum disorder (ASD) | 6,659 | 5.6 | 74.4 | 25.6 | < 0.0001 |
| Schizophrenia | 5,024 | 4.2 | 67.4 | 32.6 | < 0.0001 |
| Sleep–wake disorders | 3,830 | 3.2 | 58.1 | 41.9 | < 0.0001 |
| Bulimia nervosa | 2,946 | 2.5 | 8.9 | 91.1 | < 0.0001 |
| Suicidal behavior | 2,855 | 2.4 | 26.6 | 73.4 | < 0.0001 |
| Other psychosis | 2,201 | 1.9 | 58.3 | 41.7 | < 0.0001 |
| Bipolar disorders | 2,022 | 1.7 | 52.3 | 47.7 | < 0.0001 |
| Alcohol use disorder | 1,402 | 1.2 | 69.8 | 30.2 | < 0.0001 |
| Manic episode | 1,102 | 0.9 | 62.7 | 37.3 | < 0.0001 |
| Sexual disorders | 658 | 0.5 | 36.9 | 63.1 | < 0.0001 |
| Behavioral disorders | 365 | 0.3 | 52.3 | 47.7 | 0.004 |

**Supplementary Table S2.**
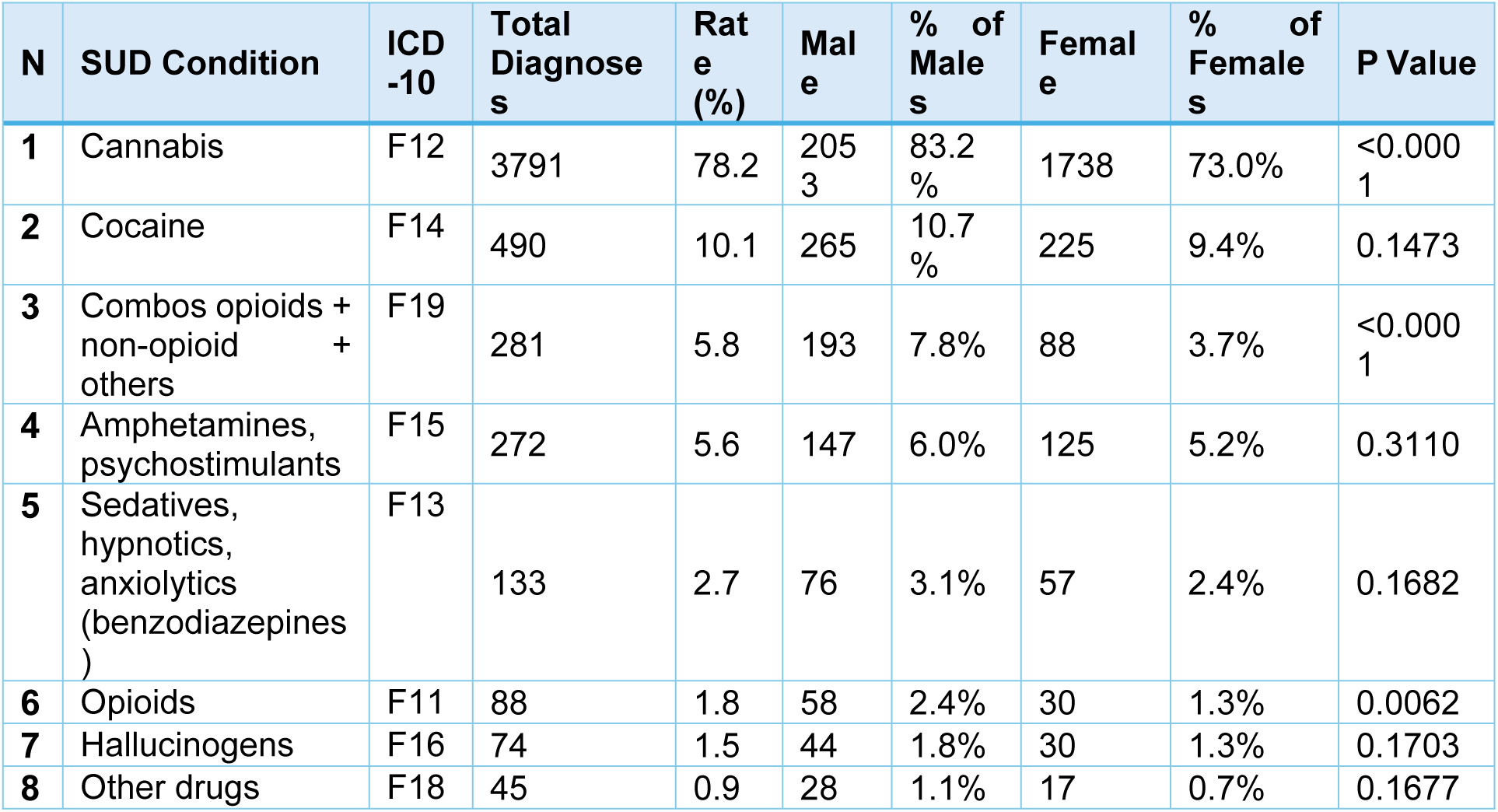
Distribution of substance use disorder (SUD) diagnoses among adolescents hospitalized in Spain between 1 January 2016 and 15 March 2020. The total number of diagnoses reflects the number of hospital records in which each SUD label was mentioned, regardless of its diagnostic position. The table includes breakdowns by sex and provides p-values derived from chi-square tests comparing male and female distributions. Rows are ordered by descending frequency (Rate %).

| N | SUD Condition | ICD -10 | Total Diagnoses | Rate (%) | Male | % of Males | Female | % of Females | P Value |
| --- | --- | --- | --- | --- | --- | --- | --- | --- | --- |
| 1 | Cannabis | F12 | 3791 | 78.2 | 2053 | 83.2% | 1738 | 73.0% | <0.0001 |
| 2 | Cocaine | F14 | 490 | 10.1 | 265 | 10.7% | 225 | 9.4% | 0.1473 |
| 3 | Combos opioids + non-opioid + others | F19 | 281 | 5.8 | 193 | 7.8% | 88 | 3.7% | <0.0001 |
| 4 | Amphetamines, psychostimulants | F15 | 272 | 5.6 | 147 | 6.0% | 125 | 5.2% | 0.3110 |
| 5 | Sedatives, hypnotics, anxiolytics (benzodiazepines) | F13 | 133 | 2.7 | 76 | 3.1% | 57 | 2.4% | 0.1682 |
| 6 | Opioids | F11 | 88 | 1.8 | 58 | 2.4% | 30 | 1.3% | 0.0062 |
| 7 | Hallucinogens | F16 | 74 | 1.5 | 44 | 1.8% | 30 | 1.3% | 0.1703 |
| 8 | Other drugs | F18 | 45 | 0.9 | 28 | 1.1% | 17 | 0.7% | 0.1677 |

**Suplemmentary Table S3.** This table summarizes the characteristics of 20 diagnostic clusters (F0-F9 for females; M0-M9 for males) derived from unsupervised analysis of ICD-10 diagnostic embeddings among Spanish adolescents hospitalised with mental health conditions. Each cluster represents a distinct data-driven subgroup identified via k-means clustering on DeBERTa-derived embeddings of diagnostic code profiles. The table includes the total number of hospitalizations in each cluster, the five most frequent ICD-10 codes within that cluster (“Top 5 ICD-10 Codes”), and the corresponding dominant semantic terms (“Top 5 Semantic Terms”), extracted using a TF-IDF-like scoring method (c-DF–IPF). Each ICD-10 code is shown alongside its matched semantic descriptor, allowing for immediate interpretation of the core diagnostic features that define each subgroup. Clusters are sex-stratified and ordered by cluster ID.

| Cluster | # Hospitalizations | Top 5 ICD-10 Codes (no. Admissions) with Semantic Terms |
| --- | --- | --- |
| **F0 (female)** | 187 | • F603 (30): borderline_personality_disorder<br>• F1210 (25): suicidal_ideations<br>• R45851 (24): personality_disorder_unspecified<br>• F609 (22): cannabis_abuse_uncomplicated<br>• F17210 (22): major_depressive_disorder_single_episode_unspecified |
| **F1 (female)** | 310 | • F17210 (96): nicotine_dependence_cigarettes_uncomplicated<br>• F1210 (92): cannabis_abuse_uncomplicated<br>• F603 (35): borderline_personality_disorder<br>• F1220 (33): conduct_disorder_unspecified<br>• F919 (30): cannabis_use_unspecified_uncomplicated |
| **F2 (female)** | 322 | • F1210 (65): borderline_personality_disorder<br>• F603 (60): cannabis_abuse_uncomplicated<br>• F17210 (54): nicotine_dependence_cigarettes_uncomplicated<br>• F1220 (33): personality_disorder_unspecified<br>• F609 (32): alcohol_abuse_uncomplicated |
| **F3 (female)** | 296 | • F603 (49): adjustment_disorder_with_mixed_disturbance_of_emotions_and_conduct<br>• F4325 (48): borderline_personality_disorder<br>• R45851 (40): suicidal_ideations<br>• F1210 (27): major_depressive_disorder_single_episode_unspecified<br>• F17210 (23): oppositional_defiant_disorder |
| **F4 (female)** | 378 | • F17210 (104): nicotine_dependence_cigarettes_uncomplicated<br>• F1210 (92): cannabis_abuse_uncomplicated<br>• F603 (65): borderline_personality_disorder<br>• F609 (37): personality_disorder_unspecified<br>• F919 (33): conduct_disorder_unspecified |
| **F5 (female)** | 161 | • F603 (26): borderline_personality_disorder<br>• F1210 (25): suicidal_ideations<br>• R45851 (19): cannabis_abuse_uncomplicated<br>• F17210 (18): conduct_disorder_adolescent-onset_type<br>• F912 (15): nicotine_dependence_unspecified_uncomplicated |
| **F6 (female)** | 210 | • F1210 (45): cannabis_abuse_uncomplicated<br>• F17210 (33): nicotine_dependence_cigarettes_uncomplicated<br>• F1220 (26): borderline_personality_disorder<br>• F603 (23): cannabis_dependence_uncomplicated<br>• F1290 (14): cannabis_abuse_with_psychotic_disorder_unspecified |
| **F7 (female)** | 170 | • F23 (49): brief_psychotic_disorder<br>• F1210 (36): unspecified_mood_affective_disorder |
|  |  | • F17210 (33): mild_intellectual_disabilities<br>• F39 (30): cannabis_abuse,_uncomplicated<br>• F70 (25): nicotine_dependence,_cigarettes,_uncomplicated |
| **F8 (female)** | 246 | • F1210 (51): cannabis_abuse,_uncomplicated<br>• F17210 (40): nicotine_dependence,_cigarettes,_uncomplicated<br>• F603 (30): borderline_personality_disorder<br>• F919 (26): conduct_disorder,_unspecified<br>• F4325 (23): adjustment_disorder_with_mixed_disturbance_of_emotions_and_conduct |
| **F9 (female)** | 102 | • F29 (91): unspecified_psychosis_not_due_to_a_substance_or_known_physiological_condition<br>• F1210 (28): cannabis_abuse,_uncomplicated<br>• F17210 (18): nicotine_dependence,_cigarettes,_uncomplicated<br>• F1220 (12): cannabis_dependence,_uncomplicated<br>• F17200 (7): deficiency_of_other_specified_b_group_vitamins |
| **M0 (male)** | 164 | • F1210 (40): cannabis_abuse,_uncomplicated<br>• F1010 (19): alcohol_abuse,_uncomplicated<br>• F17210 (17): conduct_disorder,_unspecified<br>• F919 (16): ADHD<br>• F1220 (13): nicotine_dependence,_cigarettes,_uncomplicated |
| **M1 (male)** | 316 | • F1210 (68): cannabis_abuse,_uncomplicated<br>• F17210 (39): nicotine_dependence,_cigarettes,_uncomplicated<br>• F1220 (29): conduct_disorder,_unspecified<br>• F919 (26): cannabis_dependence,_uncomplicated<br>• F12159 (24): cannabis_abuse_with_psychotic_disorder,_unspecified |
| **M2 (male)** | 334 | • F1210 (73): nicotine_dependence,_cigarettes,_uncomplicated<br>• F17210 (68): cannabis_abuse,_uncomplicated<br>• F1220 (36): conduct_disorder,_unspecified<br>• F919 (34): cannabis_dependence,_uncomplicated<br>• F1290 (27): alcohol_abuse,_uncomplicated |
| **M3 (male)** | 243 | • F1210 (31): nicotine_dependence,_cigarettes,_uncomplicated<br>• F17210 (31): conduct_disorder,_unspecified<br>• F919 (26): cannabis_abuse,_uncomplicated<br>• F17200 (18): conduct_disorder,_adolescent-onset_type<br>• F912 (18): ADHD |
| **M4 (male)** | 271 | • F1210 (61): cannabis_abuse,_uncomplicated<br>• F17210 (45): nicotine_dependence,_cigarettes,_uncomplicated<br>• F1010 (30): alcohol_abuse,_uncomplicated |
|  |  | • F1220 (27): cannabis_dependence,_uncomplicated<br>• F919 (19): other_conduct_disorders |
| **M5 (male)** | 284 | • F23 (163): brief_psychotic_disorder<br>• F1210 (81): cannabis_abuse,_uncomplicated<br>• F17210 (47): nicotine_dependence,_cigarettes,_uncomplicated<br>• F1220 (40): cannabis_dependence,_uncomplicated<br>• F70 (25): mild_intellectual_disabilities |
| **M6 (male)** | 304 | • F1210 (80): cannabis_abuse,_uncomplicated<br>• F17210 (74): nicotine_dependence,_cigarettes,_uncomplicated<br>• F1220 (34): cannabis_dependence,_uncomplicated<br>• Z818 (27): family_history_of_other_mental_and_behavioral_disorders<br>• F919 (27): conduct_disorder,_unspecified |
| **M7 (male)** | 239 | • F29 (222): unspecified_psychosis_not_due_to_a_substance_or_known_physiological_condition<br>• F1210 (87): cannabis_abuse,_uncomplicated<br>• F1220 (33): cannabis_dependence,_uncomplicated<br>• F17210 (26): cannabis_use,_unspecified,_uncomplicated<br>• F1290 (22): nicotine_dependence,_cigarettes,_uncomplicated |
| **M8 (male)** | 221 | • F1210 (40): cannabis_abuse,_uncomplicated<br>• F17210 (23): unspecified_psychosis_not_due_to_a_substance_or_known_physiological_condition<br>• F29 (23): nicotine_dependence,_cigarettes,_uncomplicated<br>• F919 (20): conduct_disorder,_unspecified<br>• F1220 (17): conduct_disorder,_adolescent-onset_type |
| **M9 (male)** | 91 | • F12159 (18): cannabis_abuse_with_psychotic_disorder,_unspecified<br>• F12259 (12): cannabis_dependence_with_psychotic_disorder,_unspecified<br>• F12959 (10): cannabis_use,_unspecified_with_psychotic_disorder,_unspecified<br>• F12151 (7): cannabis_abuse_with_psychotic_disorder_with_hallucinations<br>• F12150 (6): cannabis_abuse_with_psychotic_disorder_with_delusions |

